# Higher Blood-to-Tissue Tumor Mutational Burden Ratio Is Associated With Poorer Overall Survival in Advanced Non-Small Cell Lung Cancer

**DOI:** 10.64898/2026.08.04.26359280

**Authors:** Leeseul Kim, Jongyeop Kim, Jongwoo Kim, Seungah Yoo, Minkyu Shin, Lucas Santana Dos Santos, Young Kwang Chae

**Author notes:** **Corresponding Author:** Young Kwang Chae, MD, MPH, MBA, Northwestern Medicine Developmental Therapeutics Institute, 645 N. Michigan Avenue, Suite 1006, Chicago, IL 60611, USA. **Author Email Addresses** Leeseul Kim, Young Kwang Chae, Jongyeop Kim, Jongwoo Kim, Seungah Yoo, Minkyu Shin, Lucas Santana Dos Santos.

## Abstract

**Introduction:** Blood-based and tissue-based tumor mutational burden (bTMB and tTMB) show only moderate concordance in advanced non-small cell lung cancer (NSCLC). We hypothesized that discordance between the two measures reflects tumor heterogeneity and carries prognostic information beyond either measure alone.

**Methods:** We retrospectively identified 105 patients with advanced NSCLC who underwent both blood-based (Guardant360) and tissue-based next-generation sequencing between October 2020 and September 2024. A blood-to-tissue TMB ratio was defined as ln(1+bTMB) − ln(1+tTMB). Multivariable Cox proportional hazards models estimated associations with overall survival (OS) and progression-free survival (PFS), adjusted for age, sex, ECOG performance status, smoking history, histology, and line of therapy.

**Results:** Median follow-up was 32 months (66 deaths, 83 progression events). A higher blood-to-tissue TMB ratio was independently associated with shorter OS (HR per 1-unit increase 1.60, 95% CI 1.10–2.31; p=0.01) but not PFS (HR 1.07; p=0.67). Neither component was independently prognostic when modeled alone, whereas in a joint model ln(1+bTMB) and ln(1+tTMB) were associated with survival in opposite directions. The ratio remained associated with OS after adjustment for the highest variant allele frequency, radiographic tumor burden, and extrathoracic metastatic organ count, and was uncorrelated with each of these measures. The poorest OS occurred in the discordant High bTMB/Low tTMB group (HR 2.97, 95% CI 1.32–6.66).

**Conclusions:** Blood-to-tissue TMB discordance was independently associated with shorter overall survival, independent of ctDNA shedding and baseline disease extent. Validation in prospective cohorts with contemporaneous sampling is required.

## Introduction

Tumor mutational burden (TMB) is a genomic biomarker defined as the number of somatic mutations per megabase of interrogated coding sequence^1^. Interest in TMB arose from the observation that highly mutated tumors may generate more neoantigens and therefore be more susceptible to immune checkpoint blockade^2,3^. This concept led to broad clinical interest in tissue-derived TMB (tTMB), culminating in the tumor-agnostic accelerated approval of pembrolizumab for unresectable or metastatic TMB-high solid tumors (≥10 mut/Mb) identified by an FDA-approved assay after prior treatment failure and in the absence of satisfactory alternative treatment options^4^. However, although tTMB has become the conventional approach, it has important limitations. Tissue biopsy is invasive, often limited by specimen availability and quality, and inherently reflects only a single anatomic site at a single time point. In advanced cancer, this localized snapshot may incompletely reflect the genomic landscape of a biologically heterogeneous disease evolving across multiple metastatic sites over time^5–7^.

The development of circulating tumor DNA (ctDNA) profiling created an opportunity to assess TMB with blood samples^8^. In 2018, Gandara and colleagues reported one of the first clinically influential studies of blood-based TMB (bTMB), showing that a plasma-derived TMB assay could identify patients with previously treated NSCLC who derived progression-free survival benefit from atezolizumab^9^. Subsequent studies further supported the feasibility of bTMB as an immunotherapy biomarker^10–12^.

However, bTMB and tTMB are not interchangeable. Reported concordance is only moderate and varies with ctDNA abundance, assay design, and clinical context^9,13–16^. Because tTMB reflects a single localized specimen whereas bTMB captures tumor DNA shed from multiple disease sites, discordance between the two may carry biologic meaning, such as tumor heterogeneity, rather than reflecting measurement variability alone^9,17,18^.

Tumor heterogeneity refers to genomic and biologic diversity within and across tumor cell populations and is a fundamental feature of cancer evolution^19,20^. Tumor heterogeneity may arise from the coexistence of genetically distinct subclones within a single lesion, or from differences between anatomically separate lesions in the same patient, including primary and metastatic sites^21–23^. Greater heterogeneity has been associated with therapeutic resistance, disease progression, and adverse clinical outcomes, in part because diverse subclonal populations may enable adaptation under treatment pressure^19,22^. Several approaches have been developed to assess tumor heterogeneity, including multiregion sequencing, phylogenetic reconstruction, and quantitative genomic diversity metrics such as mutant-allele tumor heterogeneity and Shannon-type indices^17,24–27^. However, these methods are often resource-intensive, methodologically complex, and difficult to implement in routine clinical practice.

We hypothesized that discordance between bTMB and tTMB may serve as an indirect surrogate of tumor heterogeneity and be associated with survival outcomes in advanced NSCLC. Accordingly, we evaluated the blood-to-tissue TMB ratio and examined its association with overall survival and progression-free survival, including whether it provides prognostic information beyond bTMB or tTMB alone.

## Materials and Methods

### Study design and endpoints

This retrospective study was approved by the Northwestern University Institutional Review Board (STU00207117), which waived the requirement for written informed consent. The study was conducted in accordance with the Health Insurance Portability and Accountability Act and the Declaration of Helsinki, as revised in 2013.

Patients with advanced NSCLC who underwent blood-based next-generation sequencing (NGS) with available bTMB between October 2020 and September 2024 were retrospectively identified and were eligible if a corresponding tTMB result was available. When multiple tissue specimens were available, the specimen obtained closest in time to the blood-based NGS was selected. The systemic regimen initiated closest after blood collection was designated as the index regimen.

The primary endpoints were overall survival (OS) and progression-free survival (PFS), each measured from initiation of the index regimen. OS was time to death from any cause; PFS was time to radiographic or clinical progression or death, whichever occurred first. Patients without the event were censored at last follow-up or last disease assessment.

### TMB assessment

Tissue TMB was obtained from multiple NGS platforms (Tempus, PGDx, NorthShore Expanded Cancer Panel, and Massachusetts General Hospital OncoPanel). Blood TMB was uniformly derived from the Guardant360 assay. To quantify discordance while accounting for right-skewness and limiting extreme values from near-zero tTMB (Supplemental Data 1), we calculated ln(1+bTMB) − ln(1+tTMB), algebraically equivalent to ln[(1+bTMB)/(1+tTMB)], hereafter the blood-to-tissue TMB ratio.

### Assessment of ctDNA shedding and baseline disease extent

Circulating tumor DNA shedding was approximated using the highest variant allele frequency (HAF) reported by the Guardant360 assay; formal tumor fraction estimates were not available. HAF was right-skewed (Supplemental Data 2) and was log-transformed as ln(1+HAF). Radiographic tumor burden was the baseline sum of longest diameters of target lesions per RECIST v1.1, modeled per 10 mm. Metastatic organ count was the number of distinct extrathoracic organ sites involved at baseline, with pleural involvement counted separately; the primary lesion, contralateral lung, and mediastinal lymph nodes were excluded because such involvement is near-universal in advanced NSCLC.

### Statistical analysis

Continuous variables were summarized as medians with interquartile ranges and compared using Mann-Whitney U tests. Survival was estimated by the Kaplan-Meier method and compared using log-rank tests; median follow-up was estimated by reverse Kaplan-Meier. Multivariable Cox proportional hazards models estimated hazard ratios (HR) and 95% confidence intervals (CI), adjusting for age (per 10-year increment), sex, ECOG performance status, smoking history, histology, and line of therapy. Stage was excluded because of extreme imbalance (three stage III versus 102 stage IV patients).

To determine whether the association with OS was attributable to either TMB component individually, we compared a set of nested Cox models including each component alone, the ratio, and a joint model containing both; the joint model was additionally reparameterized as the ratio plus the mean of the two log-transformed values. Prespecified subgroup and sensitivity analyses, and full details of all model comparisons, are described in the Supplemental Data 3.

All analyses used R version 4.4.1 (R Foundation for Statistical Computing, Vienna, Austria). Two-sided p < 0.05 was considered significant. Data analysis was conducted from December 2025 through January 2026.

## Results

### Baseline demographic and clinical characteristics

A total of 105 patients were included (66 deaths, 83 progression events). Median follow-up was 32 months by the reverse Kaplan-Meier method. The median age was 66 years (IQR 59-73). The cohort was evenly distributed by sex (53 males [50%] and 52 females [50%]). Most patients were ever-smokers (69%), and the majority had good performance status, with 73% having an ECOG performance status of 0–1(Table 1).

**Table 1.** Baseline demographic and clinical characteristics. bTMB, blood tumor mutational burden; ECOG, Eastern Cooperative Oncology Group; ICI, immune checkpoint inhibitor; Q1, Q3, first and third quartiles; TMB, tumor mutational burden; tTMB, tissue tumor mutational burden.

| Variable | N = 105<sup>1</sup> |
| --- | --- |
| **Age** | 66 (59, 73) |
| **Sex** |  |
| Male | 53 (50%) |
| Female | 52 (50%) |
| **Smoking status** |  |
| Ever | 72 (69%) |
| Never | 33 (31%) |
| **ECOG performance status** |  |
| 2-3 | 28 (27%) |
| 0-1 | 77 (73%) |
| **Stage** |  |
| IV | 102 (97%) |
| III | 3 (3%) |
| **Histology** |  |
| Non-squamous | 85 (81%) |
| Squamous | 20 (19%) |
| **Treatment regimen** |  |
| ICI-only | 25 (24%) |
| ICI-based combination regimen | 32 (30%) |
| Targeted therapy | 38 (36%) |
| Chemotherapy | 10 (10%) |
| **Line of therapy** |  |
| ≥Second-line | 45 (43%) |
| First-line | 60 (57%) |
| **Driver alteration<sup>2</sup>** |  |
| EGFR | 18 (17%) |
| ALK | 3 (3%) |
| KRAS | 10 (10%) |
| ROS1 | 3 (3%) |
| Any driver detected | 33 (31%) |
| None detected | 72 (69%) |
| **Tissue TMB (mutations/Mb)** | 7.40 (3.70, 12.30) |
| **Blood TMB (mutations/Mb)** | 10.42 (6.70, 17.22) |
| **Log-transformed bTMB/tTMB** | 0.33 (-0.15, 0.88) |
| **bTMB/tTMB (untransformed)** | 1.46 (0.85, 2.84) |
| **Blood–treatment interval (days)** | 18 (8, 28) |
| **Blood–tissue sampling interval (days)** | 32 (13, 262) |

<sup>1</sup> n (%); Median (Q1, Q3). <sup>2</sup> Driver categories are not mutually exclusive; one patient had concurrent EGFR and ALK alterations.

Nearly all patients had stage IV disease (97%), and non-squamous histology was predominant (81%). Treatment regimens included immune checkpoint inhibitor (ICI) monotherapy (24%), ICI combined with chemotherapy and/or targeted therapy (30%), targeted therapy alone (36%), and chemotherapy alone (10%). Over half of patients received treatment in the first-line setting (57%). A driver alteration was detected in 33 patients (31%), most commonly EGFR (n=18).

The median tissue TMB was 7.40 mutations/Mb (IQR 3.70-12.30), while the median blood TMB was 10.42 mutations/Mb (IQR 6.70-17.22). The median untransformed bTMB/tTMB ratio was 1.46 (IQR 0.85-2.84), and the median log-transformed bTMB/tTMB ratio (as defined in the Methods), was 0.33 (IQR −0.15-0.88). The median interval between blood-based NGS and treatment initiation was 18 days (IQR 8-28), and the median blood–tissue sampling interval was 32 days (IQR 13-262).

Tissue biopsy sites included primary lesions (36%), metastatic lesions (29%), and lymph nodes (28%); biopsy sites were unavailable in eight patients (8%). Tissue was obtained before systemic therapy in 90 patients (86%) (Supplemental Data 4).

### Survival analysis by blood-to-tissue TMB ratio in the overall cohort across treatment regimens

Median OS decreased monotonically across tertiles of the blood-to-tissue TMB ratio (33.0, 14.0, and 9.0 months), with adjusted hazard ratios of 1.72 (95% CI 0.88-3.39) for the middle and 1.78 (95% CI 0.93-3.43) for the highest tertile versus the lowest (trend *p* = 0.09). No corresponding gradient was observed for PFS (median 7.0, 4.0, and 6.0 months; adjusted HR 1.02 and 0.94) (Figure 1A, 1B).

**Figure 1.**
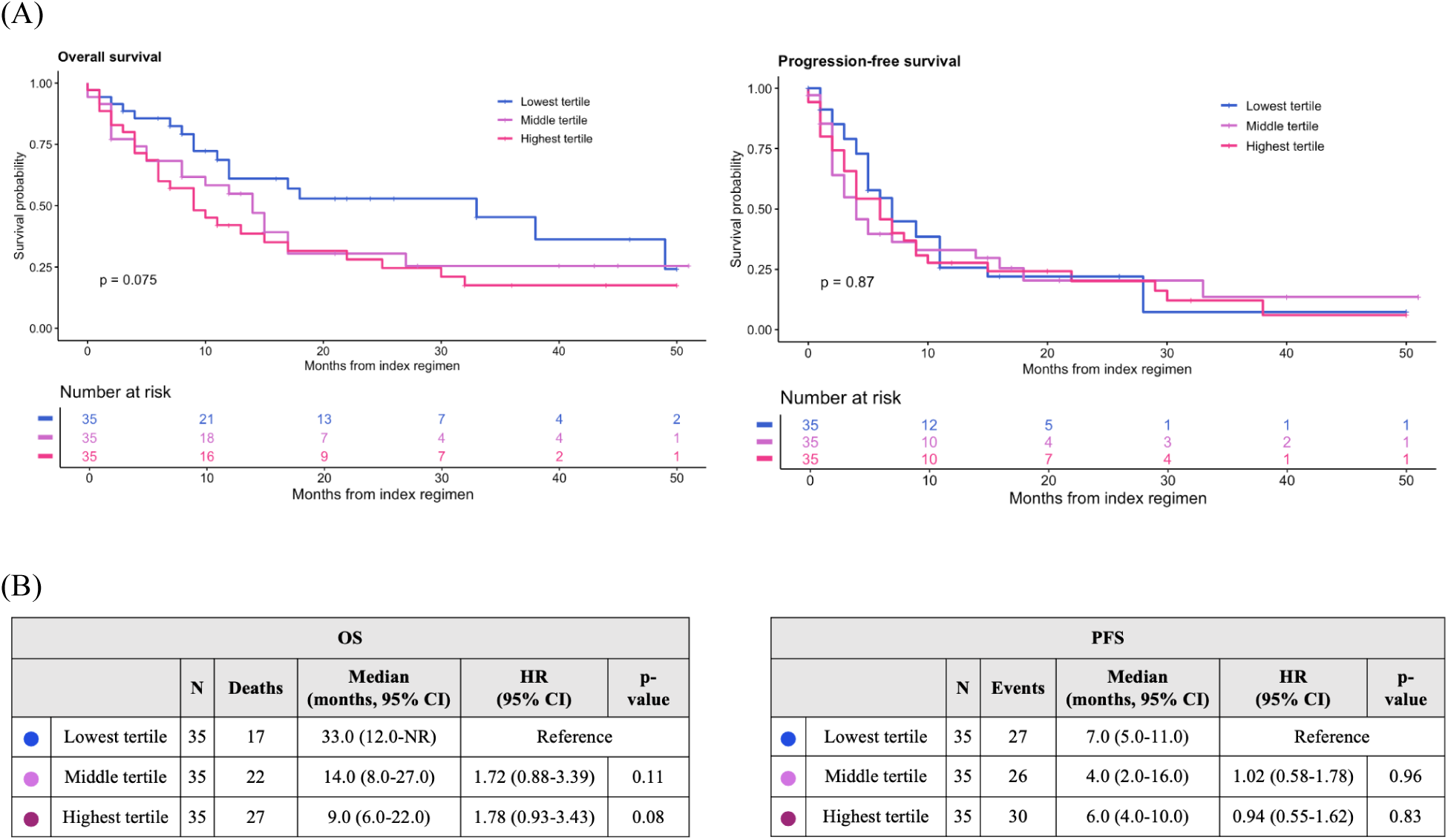
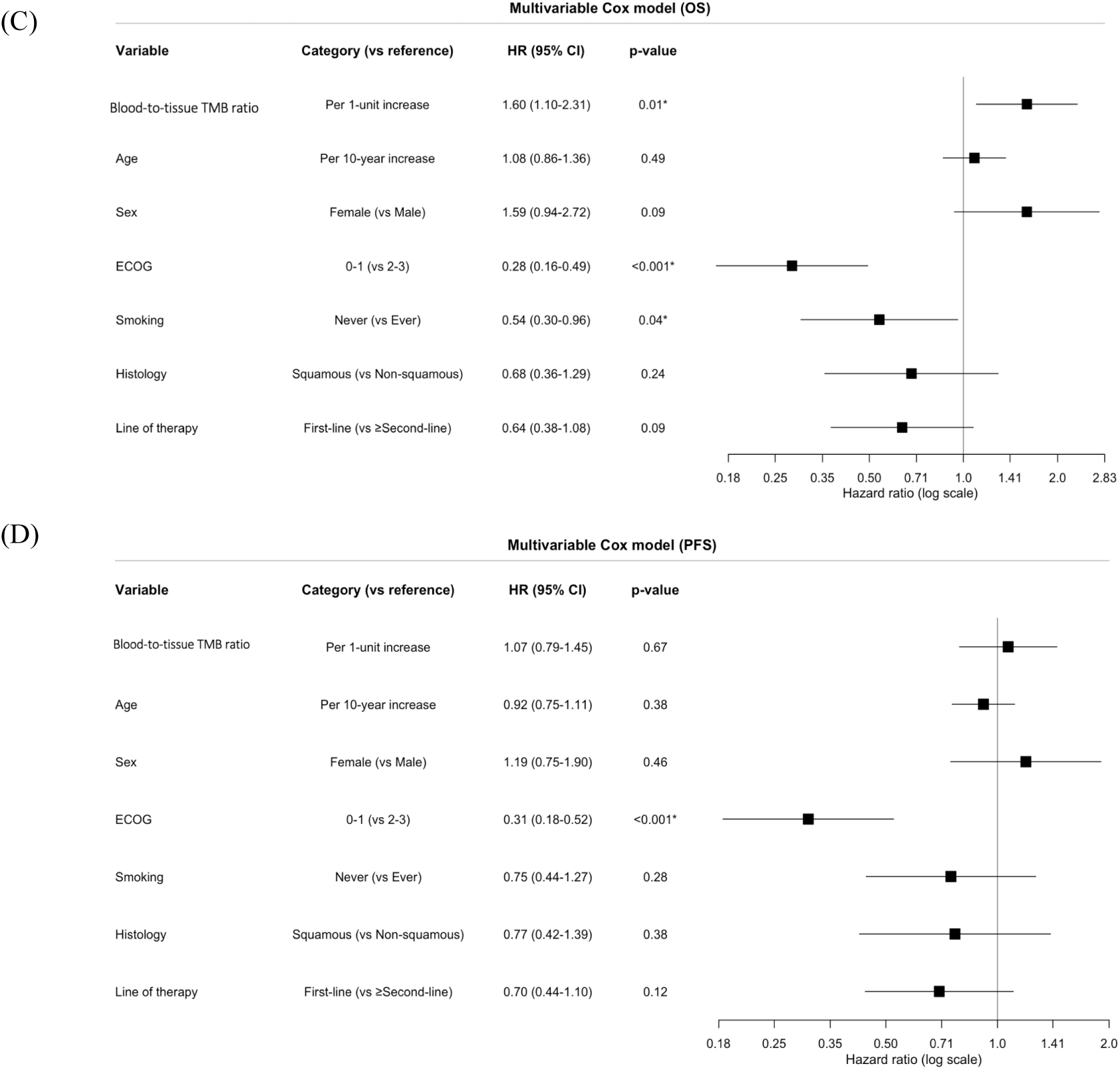
Survival by blood-to-tissue TMB ratio. (A) Kaplan-Meier curves for overall survival (left) and progression-free survival (right) by tertile of the blood-to-tissue TMB ratio. Curves are unadjusted; *p* values are from overall log-rank tests across the three tertiles. Numbers at risk are shown below each plot. (B) Median survival and hazard ratios by tertile, with the lowest tertile as the reference. Hazard ratios are from multivariable Cox proportional hazards models adjusted for age (per 10-year increment), sex, ECOG performance status, smoking history, histology, and line of therapy; median survival estimates are unadjusted. Multivariable Cox proportional hazards models for overall survival (C) and progression-free survival (D), including the blood-to-tissue TMB ratio modeled continuously per 1-unit increase together with clinical covariates. Squares indicate hazard ratios and horizontal lines 95% confidence intervals; the dashed vertical line indicates a hazard ratio of 1.CI = confidence interval; ECOG = Eastern Cooperative Oncology Group; HR = hazard ratio; NR = not reached; OS = overall survival; PFS = progression-free survival; TMB = tumor mutational burden.

When analyzed as a continuous variable, a higher blood-to-tissue TMB ratio was independently associated with shorter OS (HR per 1-unit increase 1.60, 95% CI 1.10-2.31; *p* = 0.01) but not with PFS (HR 1.07, 95% CI 0.79-1.45; *p* = 0.67) (Figure 1C, 1D).

When modeled separately, neither TMB component was significantly associated with OS (ln[1+bTMB]: HR per 1-unit increase 1.49, 95% CI 0.99-2.24; *p*=0.055; ln[1+tTMB]: HR, 0.89; 95% CI 0.62-1.26; *p*=0.50) (Supplemental Data 5). In a joint model containing both terms, higher ln(1+bTMB) was associated with shorter OS (HR 1.97; 95% CI 1.18-3.28; *p*=0.010), whereas higher ln(1+tTMB) was associated with longer OS (HR 0.67; 95% CI 0.45-0.99; *p*=0.046).

Allowing the blood and tissue TMB coefficients to vary independently did not significantly improve model fit compared with the simpler blood-to-tissue TMB ratio model (likelihood-ratio *p*=0.23). The ratio model had a slightly lower AIC than the joint model (506.96 vs 507.50) and similar optimism-corrected discrimination (C-index, 0.718 vs 0.714). In an equivalent reparameterization separating directional discordance from overall TMB magnitude, the blood-to-tissue TMB ratio remained associated with shorter OS (HR 1.72, 95% CI 1.15-2.56; *p*=0.009), whereas mean log-transformed TMB was not significantly associated with OS (HR, 1.31; 95% CI 0.85-2.04; *p*=0.22). Results for bTMB and tTMB analyzed on the untransformed scale, including dichotomized analyses using clinically validated cutoffs of 16 mutations/Mb for bTMB^9,10^ and 10 mutations/Mb for tTMB^4^, are shown in Supplemental Data 6.

### Relationship with ctDNA abundance and baseline disease extent

To determine whether the association between the blood-to-tissue TMB ratio and overall survival reflected ctDNA shedding or baseline disease extent, we examined the ratio in relation to the highest variant allele frequency (HAF) reported by the blood-based assay, the baseline sum of target lesion diameters, and the number of extrathoracic metastatic organs. HAF and extrathoracic metastatic organ count were available for all 105 patients; radiographic tumor burden was available for 101 (Supplemental Data 7).

The median HAF was 3.4% (IQR 0.9%-10.4%). The blood-to-tissue TMB ratio was not correlated with ln(1+HAF) (Spearman ρ=0.00; *p*=0.99), baseline radiographic tumor burden (ρ=0.05; *p*=0.59), or the number of extrathoracic metastatic organs (ρ=0.03; *p*=0.74). In contrast, ln(1+bTMB) was weakly correlated with ln(1+HAF) (ρ=0.24; *p*=0.01), but not with radiographic tumor burden or extrathoracic metastatic organ count(Figure 2A).

**Figure 2.**
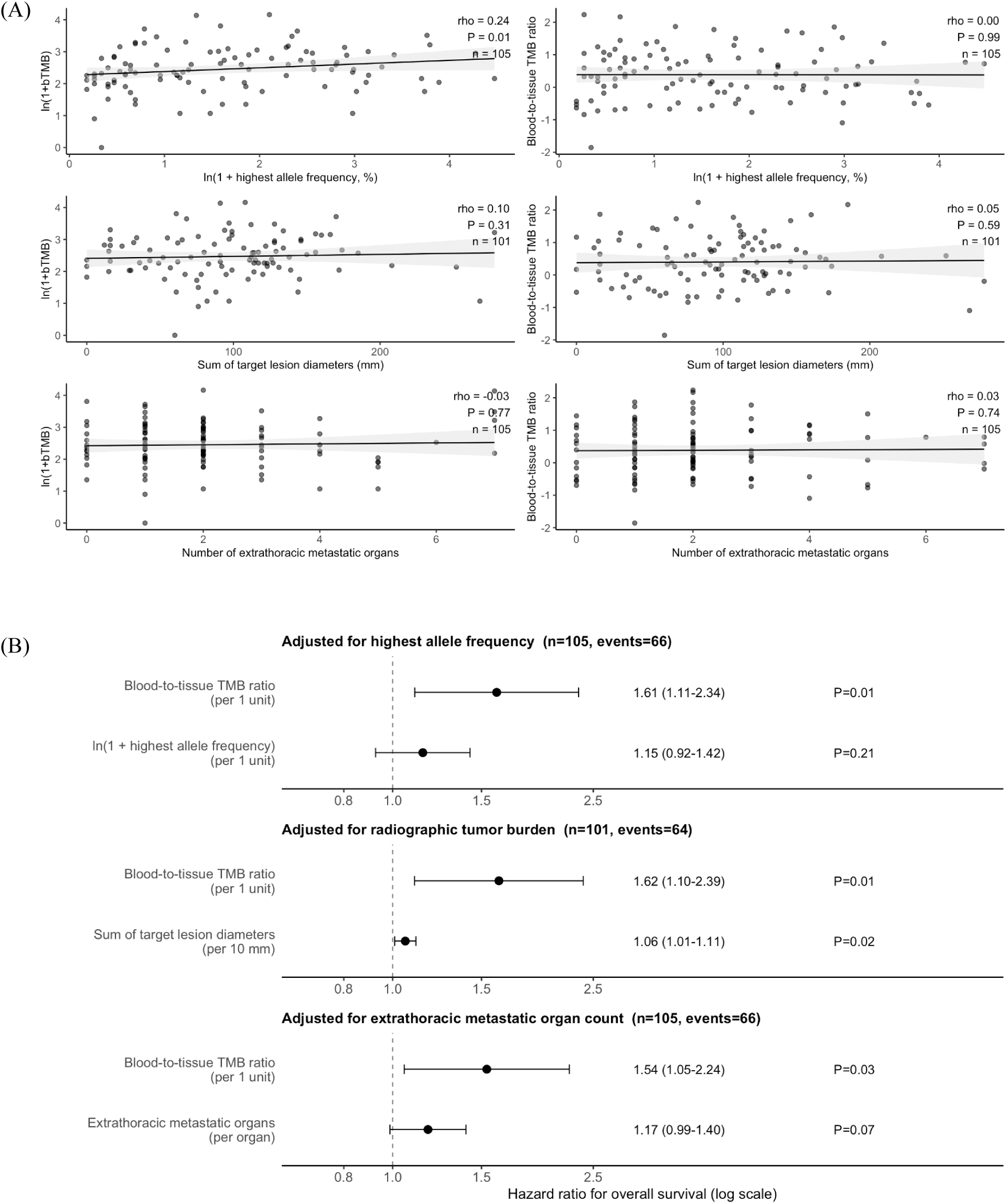
Relationship between the blood-to-tissue TMB ratio, ctDNA shedding, and baseline disease extent. (A) Scatterplots of ln(1+bTMB) (left) and the blood-to-tissue TMB ratio (right) against ln(1+HAF), sum of target lesion diameters, and extrathoracic metastatic organ count. Lines show linear fits with 95% confidence bands; Spearman rho, P values, and n are shown in each panel. (B) Forest plots of hazard ratios for overall survival from three multivariable Cox models, each containing the blood-to-tissue TMB ratio, together with ln(1+HAF), sum of target lesion diameters (per 10 mm), or metastatic organ count, adjusted for age, sex, ECOG performance status, smoking history, histology, and line of therapy. Dashed line indicates HR = 1. bTMB = blood tumor mutational burden; CI = confidence interval; ECOG = Eastern Cooperative Oncology Group; HAF = highest allele frequency; HR = hazard ratio; TMB = tumor mutational burden.

After adjustment for ln(1+HAF), the blood-to-tissue TMB log ratio remained associated with shorter OS (HR per 1-unit increase, 1.61, 95% CI 1.11-2.34; *p*=0.01), whereas ln(1+HAF) was not significantly associated with OS either in this model (HR 1.15, 95% CI 0.92-1.42; *p*=0.21)(Figure 2B)or when modeled without the ratio (HR 1.14, 95% CI 0.92-1.41; *p*=0.23). Findings were unchanged when HAF was modeled without transformation (ratio: HR 1.60, 95% CI 1.10–2.31; *p*=0.01).

The ratio also remained associated with shorter OS after adjustment for baseline radiographic tumor burden (HR 1.62, 95% CI 1.10-2.39; *p*=0.01) and extrathoracic metastatic organ count (HR 1.54, 95% CI 1.05-2.24; *p*=0.03). Greater radiographic tumor burden was independently associated with shorter OS (HR per 10-mm increase 1.06, 95% CI 1.01-1.11;*p* =0.02), whereas extrathoracic metastatic organ count showed a similar but non-significant trend (HR per organ 1.17, 95% CI 0.99-1.40; *p*=0.07)(Figure 2B).

### Four-group survival analysis stratified by bTMB and tTMB

Patients were classified using clinically validated cutoffs for bTMB and tTMB, with the Low bTMB/Low tTMB group (n=57) as the reference (Figure 3). Median OS was 15.0 months (95% CI 11.0–NR) in the reference group, 18.0 months (95% CI 14.0–NR) in Low bTMB/High tTMB (n=18), 9.0 months (95% CI 4.0–NR) in High bTMB/High tTMB (n=19), and 6.0 months (95% CI 4.0–NR) in High bTMB/Low tTMB (n=11). In adjusted analysis, the poorest OS was observed in the discordant High bTMB/Low tTMB group (HR 2.97, 95% CI 1.32-6.66; *p*< 0.01), followed by High bTMB/High tTMB (HR 2.09, 95% CI 1.07-4.08; *p* = 0.03), whereas Low bTMB/High tTMB did not differ from the reference (HR 0.98, 95% CI 0.46-2.09; *p* = 0.97). No significant differences in PFS were observed across groups.

**Figure 3.**
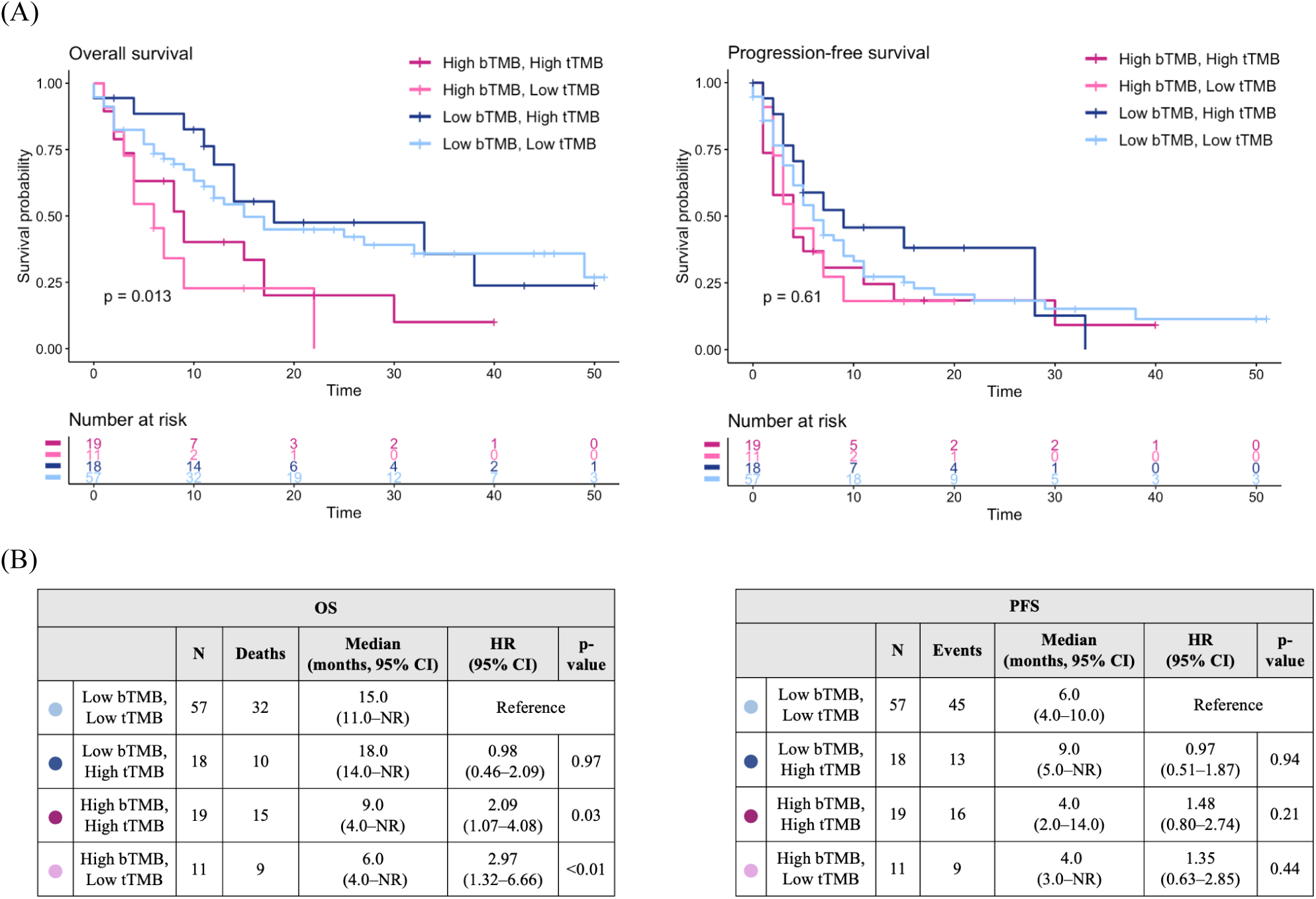
Survival by combined bTMB and tTMB status. (A) Kaplan-Meier curves for overall survival (left) and progression-free survival (right) across four groups defined by clinically validated cutoffs for bTMB (high ≥16 vs low <16 mutations/Mb) and tTMB (high ≥10 vs low <10 mutations/Mb). Curves are unadjusted; the displayed *p* value is from an overall log-rank test across all four groups. Numbers at risk are shown below each plot. (B) Median survival and hazard ratios by group. Hazard ratios are from multivariable Cox proportional hazards models adjusted for age (per 10-year increment), sex, ECOG performance status, smoking history, histology, and line of therapy, with the Low bTMB/Low tTMB group as the reference. Median survival estimates are unadjusted. bTMB = blood tumor mutational burden; CI = confidence interval; HR = hazard ratio; NR = not reached; OS = overall survival; PFS = progression-free survival; tTMB = tissue tumor mutational burden.

### Subgroup analysis by treatment line, tissue–blood time gap, regimen, and tissue biopsy site

Hazard ratios were directionally consistent with the overall cohort across all four subgroup variables (Table 2). Associations reached nominal significance among patients receiving second-line or later therapy (HR 2.08, 95% CI 1.14–3.78; *p*=0.02), those with a tissue– blood time gap at or above the median (HR 2.25, 95% CI 1.25–4.04; *p*<0.01), and those treated without ICI (HR 1.94, 95% CI 1.15-3.28; *p* = 0.01), whereas estimates in the complementary strata were of similar direction but did not reach significance (first-line, HR 1.43 [95% CI 0.90-2.25]; below-median time gap, HR 1.56 [95% CI 0.87-2.77]; ICI-containing regimen, HR 1.40 [95% CI, 0.78-2.50]). Estimates by tissue biopsy site were similar in both strata (primary lesion, HR 1.50 [95% CI 0.86-2.59]; metastatic lesion or lymph node, HR 1.58 [95% CI 0.90-2.77]). No interaction term reached statistical significance (all *p* for interaction ≥ 0.58), and confidence intervals overlapped substantially across strata, providing no evidence that the association differed by subgroup; given the limited number of events, these analyses were underpowered to detect modest effect modification and should be considered exploratory.

**Table 2.** Subgroup multivariable Cox proportional hazards analysis of blood-to-tissue TMB ratio and overall survival by treatment line, tissue-blood time gap, regimen, and tissue biopsy site. Models were adjusted for age (per 10-year increment), sex, ECOG performance status, smoking history, histology, and line of therapy where applicable, and P values for interaction are reported.

| Subgroup | Level | N | Events | HR (95% CI) | P value | P for Interaction |
| --- | --- | --- | --- | --- | --- | --- |
| **Total** |  | 105 | 66 | 1.60 (1.10-2.31) | 0.01 |  |
| **Treatment line** | First-line | 60 | 35 | 1.43 (0.90–2.25) | 0.13 | 0.75 |
|  | ≥Second-line | 45 | 31 | 2.08 (1.14–3.78) | 0.02 |  |
| **Tissue-blood time gap** | Below median | 52 | 29 | 1.56 (0.87–2.77) | 0.13 | 0.73 |
|  | Above/equal median | 53 | 37 | 2.25 (1.25–4.04) | <0.01 |  |
| **Regimen** | Non-immunotherapy | 48 | 30 | 1.94 (1.15–3.28) | 0.01 | 0.69 |
|  | Immunotherapy | 57 | 36 | 1.40 (0.78–2.50) | 0.26 |  |
| **Tissue biopsy site** | Primary lesion | 38 | 26 | 1.50 (0.86–2.59) | 0.15 | 0.58 |
|  | Metastatic lesion /Lymph node | 59 | 37 | 1.58 (0.90–2.77) | 0.11 |  |

### Blood-to-tissue TMB ratio in relation to temporal and spatial factors

To explore whether blood-tissue discordance reflected temporal evolution or spatial disease distribution, the ratio was compared across strata defined by tissue-blood sampling interval, line of therapy, and baseline disease extent (Supplemental Data 8). The ratio did not differ significantly across any stratum. Values were somewhat higher among patients with a below-median sampling interval (0.48 [IQR 0.01 to 1.09] vs 0.27 [IQR −0.42 to 0.72]; *p* = 0.08). Ratios were similar in first-line versus second-line or later settings (0.31 [IQR −0.01 to 1.00] vs 0.38 [IQR −0.18 to 0.78]; *p* = 0.56), by baseline radiographic tumor burden (0.33 [IQR −0.18 to 0.84] vs 0.44 [IQR −0.01 to 0.91]; *p* = 0.54), and by extrathoracic metastatic organ count (0.32 [IQR −0.32 to 0.89] vs 0.38 [IQR −0.03 to 0.87]; *p* = 0.63).

### Sensitivity analyses

Hazard ratios for the blood-to-tissue TMB ratio were directionally consistent across all sensitivity analyses, ranging from 1.42 to 2.17 (Supplemental Data 9). Additional adjustment for driver alteration status yielded a similar estimate (HR 1.52, 95% CI 1.00-2.30; *p* = 0.05), and restriction to the 72 patients without a detected driver alteration produced a comparable point estimate with a wider confidence interval (HR 1.42, 95% CI 0.88-2.30; *p* = 0.15). Restriction to a single tissue sequencing platform gave estimates of 2.17 (95% CI 1.26-3.73; *p* < 0.01; n = 61) and 1.75 (95% CI 0.67–4.60; *p* = 0.26; n = 41). Among the 90 patients whose tissue was obtained before systemic therapy, the association was preserved (HR 1.55, 95% CI 1.04-2.30; *p* = 0.03). Attenuation in the driver-negative and smaller-platform subsets was accompanied by substantially reduced sample size, and confidence intervals overlapped substantially with the primary estimate in all cases.

## Discussion

A higher blood-to-tissue TMB ratio, reflecting elevated bTMB relative to tTMB, was independently associated with worse overall survival in this cohort of patients with advanced NSCLC. We hypothesized that discordance between the two measures reflects tumor heterogeneity, with bTMB capturing a broader genomic landscape than tTMB derived from a single biopsy.

Our modeling analyses indicate that the prognostic signal resides in the discordance between blood-and tissue-derived TMB rather than in the absolute level of either measure. When modeled individually, neither ln(1+bTMB) nor ln(1+tTMB) was significantly associated with overall survival. In a joint model containing both terms, however, each was independently associated with survival in opposite directions, and each contributed information beyond the other. This pattern is what would be expected if the biologically relevant quantity is the difference between the two compartments rather than either value alone. In an equivalent reparameterization separating discordance from overall TMB level, the ratio remained associated with overall survival whereas mean log-transformed TMB did not. Together these findings argue that the association we observed is not simply a restatement of elevated bTMB or of high TMB in general.

When modeled on the untransformed scale, neither tTMB nor bTMB was consistently prognostic as a continuous variable. However, a bTMB threshold of ≥16 mutations/Mb, previously reported as predictive of benefit in ICI-treated populations ^9,10,28^, was associated with inferior survival in our cohort.

These findings align with prior evidence indicating that the predictive and prognostic value of TMB is highly treatment-context dependent. High tTMB has been associated with improved progression-free survival in selected ICI-only settings^29–36^, particularly with dual immune checkpoint blockade, but has not consistently translated into improved overall survival. Moreover, this predictive signal appears attenuated in chemo-immunotherapy regimens^29,37^ where cytotoxic effects and chemotherapy-induced immune modulation reduce reliance on tumor immunogenicity^38,39^. Similarly, studies of bTMB have demonstrated predictive value primarily in ICI monotherapy or ICI-only combinations^9,10,28^, with diminished utility in chemo-immunotherapy regimens and in non-ICI contexts^36,40^. Accumulating evidence suggests that elevated bTMB may also reflect adverse tumor biology. Across studies, higher bTMB has been associated with increased ctDNA tumor fraction^9,14,41^, which is itself prognostic and reflects tumor burden and systemic disease dissemination. Thus, elevated bTMB may represent not only greater mutational load, but also increased ctDNA shedding, linking it to higher disease burden and more aggressive tumor biology^42,43^.

Stratification by clinically validated bTMB and tTMB cutoffs was consistent with the continuous analysis. The poorest overall survival occurred in the discordant High bTMB/Low tTMB group, whereas the Low bTMB/High tTMB group did not differ from the reference in adjusted analysis despite the longest observed median survival. Because the High bTMB/Low tTMB group is by definition the upper extreme of the ratio, this categorical analysis represents an alternative view of the same signal rather than independent confirmation.

In our cohort, although ln(1+bTMB) was weakly correlated with ln(1+HAF), the blood-to-tissue TMB ratio showed no correlation with HAF, radiographic tumor burden, or extrathoracic metastatic organ count, and remained independently associated with shorter overall survival after separate adjustment for each of these measures. Radiographic tumor burden also remained independently associated with survival in the same models, indicating that the ratio and conventional measures of disease extent carry partly distinct prognostic information. These findings suggest that the prognostic information captured by blood-tissue TMB discordance is not solely a consequence of greater ctDNA shedding or more extensive baseline disease. Several caveats apply. HAF is an indirect proxy for ctDNA abundance rather than a direct estimate of tumor fraction. Radiographic tumor burden and metastatic organ count also incompletely characterize total disease burden, and residual confounding by ctDNA shedding, anatomic disease distribution, and unmeasured tumor volume cannot be excluded. Nonetheless, the persistence of the ratio association across these adjusted models supports its evaluation as a hypothesis-generating prognostic marker reflecting divergence in the mutational landscape sampled by blood versus tissue.

Paired tissue–plasma sequencing studies support the interpretation that bTMB-tTMB discordance may reflect biological differences captured by liquid biopsy rather than analytic variability alone^9,16^. In the paired plasma–tissue analysis of the POPLAR and OAK cohorts, Gandara et al. found that among patients with high mutation counts in both compartments, approximately one-third of variants were unique to blood and one-fourth were unique to tissue, indicating that discordance is driven in part by nonoverlapping mutational repertoires contributing to each measure^9^. Similarly, in CheckMate 848, He et al. showed that bTMB and tTMB captured overlapping but nonidentical variant sets, with the bTMB assay generally identifying more unique variants; notably, most plasma-unique variants were subclonal^15^.

This heterogeneity may be both spatial and temporal. Spatially, ctDNA is shed from multiple tumor deposits, enabling plasma-based sequencing to interrogate a broader and more heterogeneous genomic landscape than tissue sequencing derived from a single lesion^8,17,44^. In this framework, elevated bTMB relative to tTMB may reflect subclonal and spatially distributed alterations arising from unsampled deposits. Temporally, tissue and blood samples are often obtained at different time points in real-world practice, and blood-based NGS may therefore capture clonal evolution, acquired resistance, and cumulative mutation acquisition not represented in an earlier tissue specimen. Consistent with this, Gandara et al. identified longer intervals between tissue and plasma collection as a major determinant of reduced blood–tissue TMB concordance and suggested that intervening exposure to DNA-damaging therapies, such as platinum-based chemotherapy, may contribute to relative increases in bTMB^9^. Mishima et al. likewise showed that many markedly discordant cases occurred in high–tumor-fraction liquid biopsies obtained more than 3 months after tissue biopsy and often bore mutational signatures consistent with prior DNA-damaging therapy or cumulative mutation acquisition in DNA repair-deficient tumors^16^.

Our data provide limited direct support for either mechanism. The ratio did not differ significantly by sampling interval and, if anything, was somewhat higher among patients sampled at shorter intervals (p = 0.08), a direction opposite to that expected if interval clonal evolution were the principal driver. Nor was the ratio correlated with the number of extrathoracic metastatic organs or different between organ-count strata.

A notable pattern in our data is that the prognostic association was confined to overall survival; no relationship with progression-free survival was observed. It may reflect the ratio capturing a feature of tumor biology that manifests over the full disease course, through the emergence of resistant subclones under sequential treatment pressure, diminished tolerance of subsequent therapy, or a broader propensity toward aggressive behavior not reflected in short-term treatment response.

This study has several limitations. First, blood and tissue samples were not collected in a matched prospective manner, introducing temporal variability between bTMB and tTMB measurements. Second, this was a retrospective cohort with heterogeneous treatment regimens and lines of therapy, limiting our ability to isolate treatment-specific effects on survival. Third, tissue-based TMB assays were not uniform across patients, introducing assay-level heterogeneity in tTMB measurement. Fourth, ctDNA shedding was approximated using the highest variant allele frequency reported by the blood-based assay rather than a direct estimate of tumor fraction, and residual confounding by shedding cannot be fully excluded. Finally, this was a small, single-institution study, which limits statistical power and generalizability.

In conclusion, a higher blood-to-tissue TMB log ratio was independently associated with shorter overall survival after adjustment for clinicopathologic characteristics and baseline disease extent. This directional blood–tissue TMB metric may capture prognostically relevant differences not consistently reflected by either TMB measure when evaluated separately. However, whether these differences primarily represent tumor heterogeneity, temporal evolution, assay-related variation, or other biological factors requires validation in prospectively collected cohorts with contemporaneous and analytically harmonized blood and tissue testing.

## Data Availability

All data produced in the present study are available upon reasonable request to the authors

## Declarations

### Ethics Approval and Consent to Participate

This study was approved by the Institutional Review Board of Northwestern University Feinberg School of Medicine (STU00207117). The study was conducted in accordance with applicable institutional and ethical standards.

### Consent for Publication

This retrospective study used de-identified patient data; therefore, consent for publication was not required.

### Declaration of Interests

Young Kwang Chae reports research funding from AbbVie, Bristol Myers Squibb, Biodesix, Freenome, Predicine, Tempus, Imagene AI, Picture Health, OncoHost, and Regeneron; consulting or advisory roles with Roche/Genentech, AstraZeneca, Foundation Medicine, NeoGenomics, Guardant Health, Boehringer Ingelheim, Biodesix, ImmuneOncia, Lilly Oncology, Merck, Takeda, Lunit, Jazz Pharmaceuticals, Tempus, Bristol Myers Squibb, Regeneron, NeoImmuneTech, Eisai, OncoHost, Novocure, Picture Health, GeneCker, and Bayer; and speaker engagements with Guardant Health, Natera, Novocure, AstraZeneca, Regeneron, AbbVie, Bristol Myers Squibb, Merck, and Lilly Oncology. These relationships were unrelated to the present study.

The remaining authors declare no competing interests.

### Funding

This research did not receive any specific grant from funding agencies in the public, commercial, or not-for-profit sectors.

### Declaration of Generative AI and AI-Assisted Technologies in the Writing Process

During the preparation of this manuscript, the authors used ChatGPT (OpenAI) and Claude (Anthropic) to improve the language and readability of the manuscript and to assist with statistical code and analysis planning. All analyses were reviewed and verified by the authors, who take full responsibility for the content of the publication.

## List of Supplementary Material

Supplemental Data 1. docx

## Notes

### Author Declarations

This retrospective study was approved by the Northwestern University Institutional Review Board (STU00207117). The requirement for written informed consent was waived by the institutional review board.

