## Supplementary for "Higher Blood-to-Tissue Tumor Mutational Burden Ratio Is Associated With Poorer Overall Survival in Advanced Non-Small Cell Lung Cancer"

Supplemental Data 1. Distribution of the blood-to-tissue TMB ratio
Supplemental Data 2. Distribution of highest allele frequency
Supplemental Data 3. Supplementary Methods
Supplemental Data 4. Tissue biopsy site, sequencing platform, and timing
Supplemental Data 5. Comparison of TMB parameterizations
Supplemental Data 6. Blood and tissue TMB analyzed individually
Supplemental Data 7. Baseline ctDNA shedding and disease extent measures
Supplemental Data 8. Blood-to-tissue TMB ratio across temporal and spatial strata
Supplemental Data 9. Sensitivity analyses

Supplemental Data 1. (A) Distribution of bTMB/tTMB. (B) Distribution of log-transformed blood-to-tissue TMB ratio.

(A)

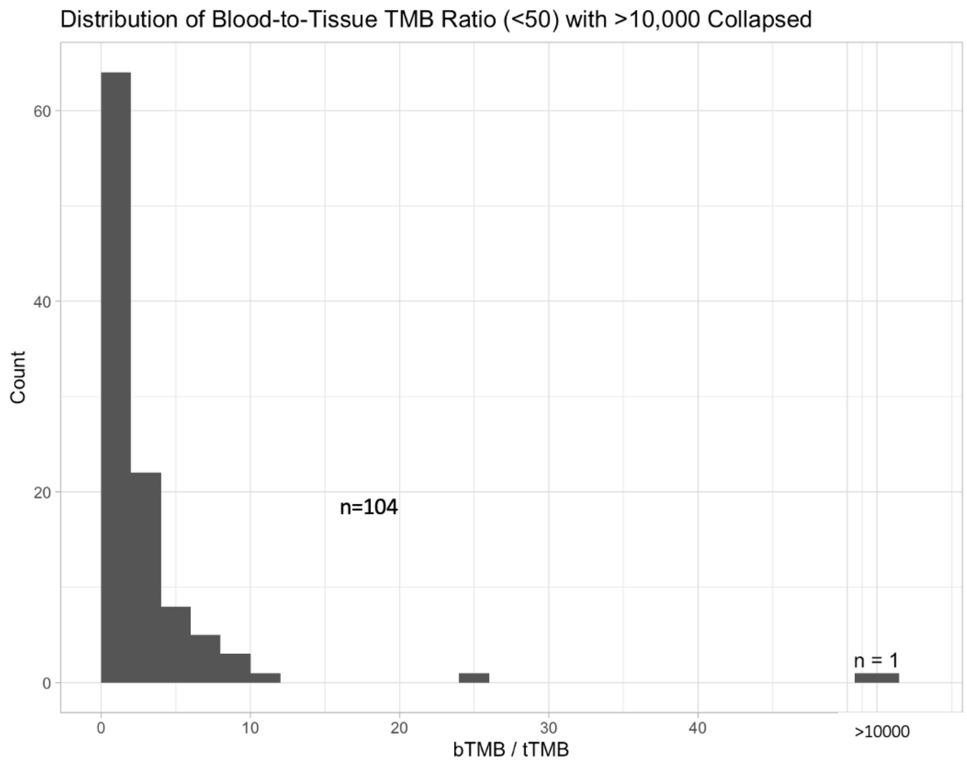


(B)

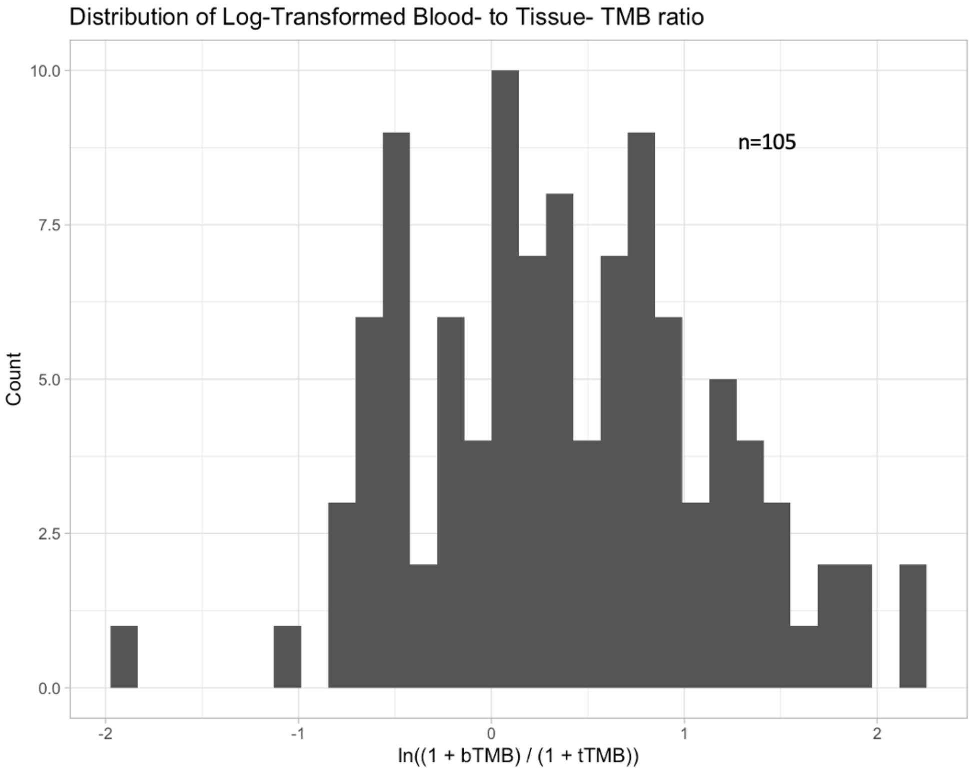

Supplemental Data 2. (A) Distribution of highest allele frequency (B) Distribution of log-transformed highest allele frequency.

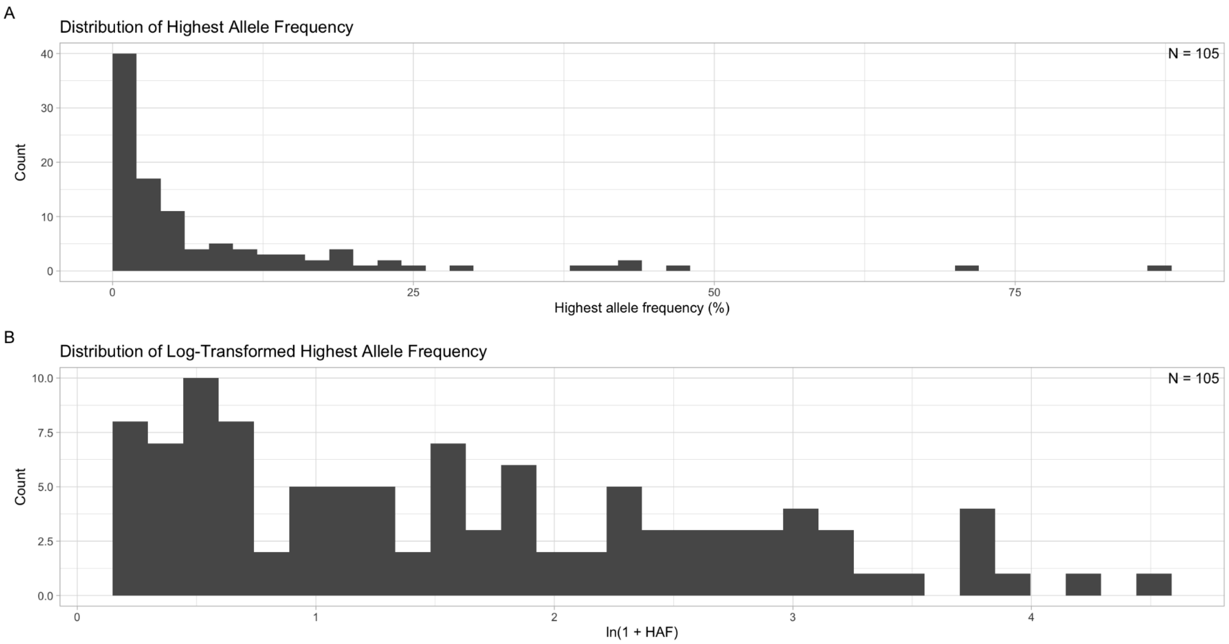

**Supplemental Data 3.**

**Comparison of TMB parameterizations**

Models were fitted on the identical complete-case sample and adjusted for the same covariates: clinical covariates alone, covariates plus ln(1+bTMB), covariates plus ln(1+tTMB), covariates plus the blood-to-tissue TMB ratio, and a joint model containing both ln(1+bTMB) and ln(1+tTMB). Because the ratio is the difference between the two log-transformed components, the ratio model is nested within the joint model under the constraint that the component coefficients are equal in magnitude and opposite in direction; this constraint was tested by likelihood ratio test. The joint model was additionally reparameterized as the ratio plus the mean of the two log-transformed values, an algebraically equivalent formulation distinguishing blood-tissue discordance from overall TMB level. Model fit was compared using log-likelihood and the Akaike information criterion, with lower values indicating better fit. Discrimination was assessed using Harrell's C-index, corrected for optimism by bootstrap resampling with 1000 replicates using the rms package. Correlations between log-transformed TMB terms were assessed using the Pearson coefficient, as these variables enter the models as linear terms.

**Four-group analysis by bTMB and tTMB**

Patients were classified into four groups using clinically validated cutoffs of 16 mutations/Mb for bTMB and 10 mutations/Mb for tTMB. The Low bTMB/Low tTMB group was designated as the reference because it was the largest stratum. Survival was compared using Kaplan-Meier estimates and multivariable Cox models adjusted for the same covariates as the primary analysis.

**Subgroup analysis**

Subgroup analyses evaluated the association between the blood-to-tissue TMB ratio and OS within strata defined by treatment line (first-line versus second-line or later), tissue-blood sampling interval (below versus at or above the median), index regimen (immunotherapy-containing versus non-immunotherapy), and tissue biopsy site (primary lesion versus metastatic lesion or lymph node). Within each stratum, a separate multivariable Cox model estimated the hazard ratio per 1-unit increase in the ratio, adjusted for the standard covariates; line of therapy was omitted from models stratified by treatment line. Analyses were restricted to patients with non-missing data for the relevant subgroup variable. Effect modification was assessed using models including the subgroup variable, the ratio, their multiplicative interaction term, and the same covariates, with Wald p values reported for interaction terms.

**Blood-to-tissue TMB ratio in relation to temporal and spatial factors**

The ratio was compared across strata defined by tissue-blood sampling interval, line of therapy, baseline radiographic tumor burden, and extrathoracic metastatic organ count, each dichotomized at the cohort median except line of therapy, using two-sided Mann-Whitney U tests.

**Sensitivity analyses**

Because oncogene-driven tumors may have systematically lower tissue TMB, models were fitted with additional adjustment for driver alteration status (any detected EGFR, ALK, KRAS, or ROS1 alteration) and separately restricted to patients without a detected driver alteration. To address heterogeneity in tissue sequencing platforms, analyses were restricted to each of the two most frequently used assays. To address potential effects of intervening systemic therapy on tissue mutational profiles, analyses were restricted to patients whose tissue was obtained before initiation of systemic therapy. Each model retained the same adjustment covariates as the primary analysis.

Supplemental Data 4. Tissue biopsy site, sequencing platform, and timing relative to systemic therapy. MGH = Massachusetts General Hospital; NGS = next-generation sequencing.

| **Tissue biopsy site** | **N (%)** |
|---|---|
| Primary lesion | 38 (36%) |
| Metastatic lesion | 30 (29%) |
| Lymph node | 29 (28%) |
| Unknown | 8 (8%) |
| **Tissue NGS platform** | |
| Tempus | 61 (58%) |
| PGDx | 41 (39%) |
| NorthShore Expanded Cancer NGS Panel | 2 (2%) |
| ONCOPANEL (MGH) | 1 (1%) |
| **Tissue biopsy timing** | |
| Before systemic therapy | 90 (86%) |
| After systemic therapy | 15 (14%) |

Supplemental Data 5. Comparison of tumor mutational burden parameterizations in multivariable Cox models for overall survival. Each row is a separate multivariable Cox model fitted on the identical complete-case sample (N = 105; 66 deaths), adjusted for age (per 10-year increment), sex, ECOG performance status, smoking history, histology, and line of therapy. Hazard ratios are per 1-unit increase. Ratio = ln(1+bTMB) − ln(1+tTMB); mean TMB = [ln(1+bTMB) + ln(1+tTMB)]/2. The two-term models are algebraically equivalent reparameterizations and therefore share identical log-likelihood, AIC, and C-index; in these models, estimates are presented in the order listed in the Model column. Lower AIC indicates better model fit. C-index corrected for optimism by bootstrap resampling (1000 replicates). AIC = Akaike information criterion; bTMB = blood tumor mutational burden; CI = confidence interval; df = degrees of freedom; ECOG = Eastern Cooperative Oncology Group; HR = hazard ratio; TMB = tumor mutational burden; tTMB = tissue tumor mutational burden.

| Model | TMB df | HR (95% CI) | P value | Log-likelihood | AIC | C-index, apparent | C-index, corrected |
|---|---|---|---|---|---|---|---|
| Covariates only | 0 | — | — | -249.50 | 510.99 | 0.737 | 0.711 |
| ln(1+bTMB) alone | 1 | 1.49 (0.99-2.24) | 0.055 | -247.67 | 509.34 | 0.737 | 0.706 |
| ln(1+tTMB) alone | 1 | 0.89 (0.62-1.26) | 0.501 | -249.27 | 512.54 | 0.741 | 0.710 |
| Ratio | 1 | 1.60 (1.10-2.31) | 0.014 | -246.48 | 506.96 | 0.745 | 0.718 |
| ln(1+bTMB) + ln(1+tTMB) | 2 | 1.97 (1.18-3.28); 0.67 (0.45-0.99) | 0.010; 0.046 | -245.75 | 507.50 | 0.748 | 0.714 |
| Ratio + mean TMB | 2 | 1.72 (1.15-2.56); 1.31 (0.85-2.04) | 0.009; 0.222 | -245.75 | 507.50 | 0.748 | 0.714 |

Supplemental Data 6. Association of (A) blood and (B) tissue TMB with overall survival and progression-free survival across total, ICI-treated, and non-ICI treated, derived from multivariable-adjusted Cox survival models adjusting for age, sex, ECOG performance status, smoking history, histology, and line of therapy.

(A)

| Analysis group | bTMB | Endpoint | HR (95% CI) | P-value |
|---|---|---|---|---|
| Total | Per 1 mutation/Mb | OS | 1.01 (0.99–1.04) | 0.18 |
| Total | Per 1 mutation/Mb | PFS | 1.02 (0.99–1.04) | 0.18 |
| Total | ≥16 vs <16 | OS | 2.35 (1.34–4.10) | <0.01 |
| Total | ≥16 vs <16 | PFS | 1.44 (0.87–2.38) | 0.16 |
| ICI treatment | Per 1 mutation/Mb | OS | 1.01 (0.98–1.04) | 0.55 |
| ICI treatment | Per 1 mutation/Mb | PFS | 1.01 (0.98–1.04) | 0.55 |
| ICI treatment | ≥16 vs <16 | OS | 2.41 (1.10–5.25) | 0.03 |
| ICI treatment | ≥16 vs <16 | PFS | 1.64 (0.80–3.39) | 0.18 |
| Non-ICI treatment | Per 1 mutation/Mb | OS | 1.02 (0.99–1.05) | 0.22 |
| Non-ICI treatment | Per 1 mutation/Mb | PFS | 1.04 (1.01–1.08) | 0.02 |
| Non-ICI treatment | ≥16 vs <16 | OS | 2.22 (0.94–5.24) | 0.07 |
| Non-ICI treatment | ≥16 vs <16 | PFS | 1.72 (0.79–3.74) | 0.17 |

(B)

| Analysis group | tTMB | Endpoint | HR (95% CI) | P-value |
|---|---|---|---|---|
| Total | Per 1 mutation/Mb | OS | 1.00 (0.97-1.03) | 0.95 |
| Total | Per 1 mutation/Mb | PFS | 1.01 (0.98-1.05) | 0.39 |
| Total | ≥10 vs <10 | OS | 1.20 (0.70–2.04) | 0.51 |
| Total | ≥10 vs <10 | PFS | 1.15 (0.70–1.87) | 0.58 |
| ICI treatment | Per 1 mutation/Mb | OS | 1.00 (0.95–1.06) | 0.99 |
| ICI treatment | Per 1 mutation/Mb | PFS | 1.02 (0.96–1.07) | 0.57 |
| ICI treatment | ≥10 vs <10 | OS | 1.40 (0.64–3.07) | 0.41 |
| ICI treatment | ≥10 vs <10 | PFS | 1.17 (0.53–2.57) | 0.70 |
| Non-ICI treatment | Per 1 mutation/Mb | OS | 0.99 (0.95–1.04) | 0.77 |
| Non-ICI treatment | Per 1 mutation/Mb | PFS | 1.02 (0.98–1.06) | 0.32 |
| Non-ICI treatment | ≥10 vs <10 | OS | 0.76 (0.34–1.68) | 0.50 |
| Non-ICI treatment | ≥10 vs <10 | PFS | 1.37 (0.69–2.69) | 0.37 |

Supplemental Data 7. Baseline ctDNA shedding and disease extent measures.[1] Median (Q1, Q3). [2] Available for 101

| Variable | N = 105[1] |
|---|---|
| Highest allele frequency (%) | 3.40 (0.90, 10.40) |
| Sum of target lesion diameters (mm)[2] | 97 (61, 128) |
| Extrathoracic metastatic organs | 2 (1, 3) |

Supplemental Data 8. Boxplots of blood-to-tissue TMB ratio according to tissue-blood time gap (A), line of therapy (B), radiographic tumor burden (C), and number of extrathoracic metastatic organs (D). All strata were dichotomized at the cohort median except the line of therapy. P values were calculated using the Wilcoxon rank-sum test.

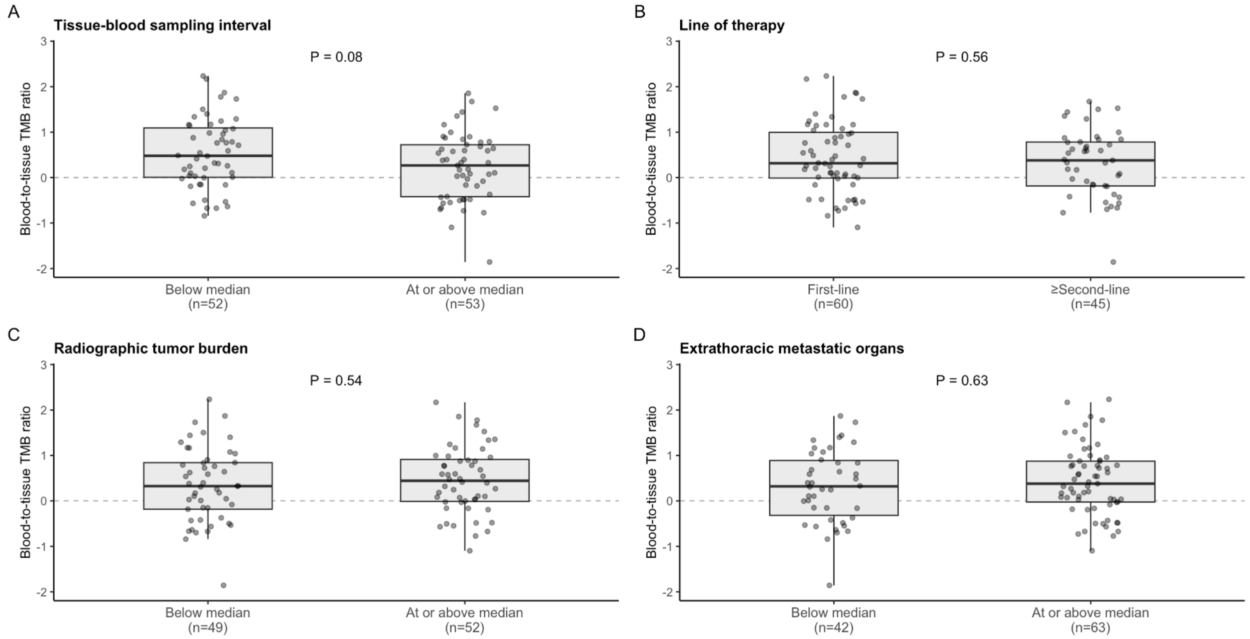

Supplemental Data 9. Sensitivity analyses of the blood-to-tissue TMB ratio and overall survival. Each row represents a separate multivariable Cox proportional hazards model adjusted for age (per 10-year increment), sex, ECOG performance status, smoking history, histology, and line of therapy. Hazard ratios are per 1-unit increase in the blood-to-tissue TMB ratio. Driver alterations comprised EGFR, ALK, KRAS, and ROS1. CI, confidence interval; ECOG, Eastern Cooperative Oncology Group; HR, hazard ratio; PGDx, Personal Genome Diagnostics; TMB, tumor mutational burden.

| Analysis | N | Events | HR (95% CI) | P value |
|---|---|---|---|---|
| Full cohort (primary analysis) | 105 | 66 | 1.60 (1.10-2.31) | 0.01 |
| Adjusted for driver alteration status | 105 | 66 | 1.52 (1.00-2.30) | 0.05 |
| Driver-negative patients only | 72 | 48 | 1.42 (0.88-2.30) | 0.15 |
| Tissue TMB from Tempus platform only | 61 | 40 | 2.17 (1.26-3.73) | <0.01 |
| Tissue TMB from PGDx platform only | 41 | 24 | 1.75 (0.67-4.60) | 0.26 |
| Pre-treatment tissue biopsy only | 90 | 56 | 1.55 (1.04-2.30) | 0.03 |
